# Relating Standardized Automated Perimetry Performed with Stimulus Sizes III and V in Eyes With Field Loss due to Glaucoma and NAION

**DOI:** 10.1101/2024.08.01.24311376

**Authors:** David Szanto, Michael Wall, Luke X Chong, Mark J Kupersmith

## Abstract

**Objective:** Standard automated perimetry (SAP) visual field (VF) results are more repeatable using Goldmann stimulus size V (stimV) in eyes with moderate/severe deficits due to glaucoma. There are few reports relating VFs using stimulus size V and III, typically used in the clinic for glaucoma, and none for non-arteritic anterior ischemic optic neuropathy (NAION). We hypothesized that we could compare and relate the VFs with both stimuli for glaucoma and NAION.

**Methods:** We utilized 1992 same-day pairs of stimIII and stimV SAP VFs using the 24-2 strategy for eyes with glaucoma or NAION. We explored the optimal threshold to censor the raw sensitivities, prior to calculating age-standardized total deviations (TD). We determined the mean and standard deviation of the differences among all TD pairs. We computed a line of best fit to determine closeness to the line of unity.

**Results:** The ideal censoring conversion threshold was 21 dB for stimIII and 24 dB for stimV. The difference between stimV and stimIII censored (0.0 ± 1.9 dB) and uncensored (0.4 ± 2.6 dB) TD pairings strongly correlate with each other (r^2^ = 0.70, p < 0.001). The line of best fit from these pairings has a slope of 0.92, which is similar to that of the line of unity (m = 1).

**Conclusion:** Censoring plus age correction is a valid method of comparison between stimIII and stimV SAP VFs with moderate to severe VF loss due to optic nerve disorders.

**Translational Relevance:** StimIII and stimV TDs are interchangeable in clinical practice.

## Introduction

Standard Automated Perimetry (SAP) is the most common method used for VF testing to measure visual sensitivities at multiple points to detect deficits in the central 30 degrees of vision. The Goldmann stimulus size III (stimIII) is widely used but subject to useful dynamic range limitation in eyes that have moderate to severe deficits.^1^ The stimulus size V (stimV), which uses a larger stimulus, has a wider useful dynamic range and is more reliable for testing eyes with more advanced glaucoma VF loss and also has better test-retest repeatability.^2^

A key challenge is comparing or converting visual field (VF) results obtained with different stimulus sizes, particularly for longitudinal data or inter-patient comparisons. Converting VFs obtained with stimV to one that would have been obtained with stimIII can standardize data, making it easier to track disease progression and compare results across studies and clinical settings. However, this conversion process has not been standardized due to the differences in sensitivity and response characteristics between the two stimulus sizes.

Censoring sensitivities below a certain threshold is necessary because data points below about 20 dB levels are dominated by noise, making them unreliable and unrepeatable.^3^ Within this range, there is high retest variability which distorts statistical measures and obscures meaningful patterns. Censoring involves adjusting all sensitivity values below a specific threshold to that threshold value. While the exact cutoff for the useful dynamic range is debated (somewhere below between 17 and 25 dB), setting these low sensitivity values in both stimuli to a predefined threshold should minimize variability with more consistent and interpretable data.^4^

Glaucoma and non-arteritic anterior ischemic optic neuropathy (NAION) are both leading causes of vision impairment in adults.^5,6^ Both cause irreversible vision loss ranging from mild visual field defects to blindness, and can significantly impact the quality of life; they are monitored using VF testing.^5,6^ Since many patients with both diseases have severely depressed VFs, stimV could be used to better monitor the disease progression. This would be particularly helpful in clinical trials that investigate therapies for eyes with moderate to severe VF deficits. Recently, new methods to analyze patterns of VF loss have complemented more global measures of VF loss. Quantifying the specific losses in regions of interest should lead to more precise monitoring and gauging the effects of therapy. Relating these patterns for stimV and stimIII in the same patient or participant is needed. However, we currently cannot directly compare stimV VFs to stimIII VFs, particularly as stimIII VFs are more likely used earlier in the disease course for glaucoma when it is mild.

This study explored whether we could determine a reasonable threshold censoring level for each point in the 24-2 VF and conversion factor of stimV to stim III VFs in individuals with moderate to severe VF loss due to glaucoma and NAION. We used two existing datasets that contained both stimIII and stimV data for the same individuals.

## Methods

This study was approved by the Institutional Review Board of the Icahn School of Medicine at Mount Sinai and required no additional consent as the data used were de-identified and derived from participants who had consented for use of their data at multiple study institutions. The study was conducted according to the tenets of the Declaration of Helsinki.

### Study Design and Participants

#### Glaucoma

Glaucoma stimIII and stimV data were received from a trial investigating differences in variability between differently sized perimetric stimuli and their ability to discriminate between healthy and damaged visual fields in glaucoma patients. The study compared abnormal test locations across different stimuli sizes to compare findings and extend the analysis to size modulation perimetry. This study involved data (previously reported) from 120 participants with glaucoma with moderate to severe VF loss who underwent same-day VF testing using both 24-2 SITA-Standard size III and full threshold size V at the University of Iowa Department of Ophthalmology and Visual Sciences.^7^ Participants were included if they had glaucomatous optic disc changes with abnormal conventional automated perimetry and were diagnosed with primary, secondary, or normal-tension glaucoma with no other vision-affecting diseases. Exclusion criteria included cataract causing visual acuity worse than 20/30, pupil size less than 2.5 mm, age under 19, or being pregnant at the time of study entry.

#### NAION

NAION data were received from the Quark207 trial, a multinational, prospective, five-armed randomized controlled trial aimed at investigating the safety and efficacy of a biologic in individuals, ages 50–80, diagnosed with NAION within 14 days of vision loss, meeting study entry criteria.^8^ The study included 729 participants, who were using the Humphrey Field Analyzer with the 24-2 SITA-Standard size III and full threshold size V, which was added after recruitment began. The two stimulus types were tested on the same day. Raw sensitivity values in decibels (dB) were recorded for each test. VFs were measured at screening, day 1 of enrollment, two months, six months, and up to one year. There were same-day VFs using both stimuli for participants at month 6 (493), month 12 (414), and various unscheduled times (32).

##### Data Censoring and Conversion

We determined optimal censoring thresholds by comparing the average difference of censored TD values between the results for both stimuli and selecting the thresholds for each stimuli that minimized this difference. We then censored sensitivities by replacing values below the defined threshold with that value. We then converted the sensitivities to age-corrected total deviation (TDs) values using a normative database for both stimuli. We also determined the optimal censoring thresholds for each disease separately to demonstrate that the data can be effectively combined in a threshold analysis.

##### Data Visualization & Analysis

We performed all statistical analyses in a Jupyter notebook with Python 3.8.8. All visualizations were done with the open-source python module “matplotlib”.^9^ We plotted pairs of TDs for all points in all VFs (103,584 pairings) as well as the line of unity (y = x) showing the perfect agreement of points. We also plotted the line of best fit and computed a coefficient of determination. We calculated mean and standard deviations between pairs of TDs, focusing on both uncensored and censored pairs (where at least one pairing is censored) by condition. We performed a paired t-test on the difference of all TD pairings.

## Results

We investigated a total of 1992 VFs from 706 participants where 120 had moderate to severe glaucoma and 586 had NAION. In participants with glaucoma, the mean age was 67.8 ± 9.3 years and 39% were male, and for participants with NAION the mean age was 61.3 ± 7.7 years and 69% were male (Table 1). The mean TD after censoring in participants with glaucoma for stimIII and stimV was −4.7 ± 3.7 dB and −4.2 ± 3.9 dB respectively, and for NAION was −7.2 ± 3.4 dB and −7.3 ± 3.8 dB respectively. We discovered that the optimal censoring threshold was 21 for stimIII and 24 for stimV, which provided the lowest mean difference and low standard deviation (Figure 1, Supplemental Figure 1).

Overall, the average difference between stimV and stimIII TD pairings for uncensored pairs was 0.4 ± 2.6 dB, and a censored pair difference of 0.0 ± 1.9 dB (Figure 2). A paired t-test analyzing differences of TDs show a statistically significant nonzero (0.1 dB) difference between the two stimuli (p < 0.001). Glaucoma VF differences between stimIII and stimV were remarkably consistent over all subtypes, showing an uncensored pair difference of 0.4 ± 2.6 dB and a censored pair difference of 0.0 ± 1.9 dB. NAION VFs consist of an uncensored pair difference of 0.2 ± 2.9 dB and a censored pair difference of −0.3 ± 1.8 dB. Notably, 63% of pairings in NAION were censored. We plotted a line of best fit along all pairs of points, and we found that it was similar to the line of unity (Figure 3). There was a strong correlation between TD pairings (r^2^ = 0.70). Opting to not censor TD pairings reveals that, beyond a certain threshold, data points increasingly deviate from the line of unity in an unpredictable way (Figure 4).

In performing threshold analysis on just glaucoma, we found that the optimal censoring threshold was 20 dB for stimIII and 22 dB for stimV. Using these thresholds for glaucoma TD pairings, the resultant line of best fit had a slope of 0.91 with strong correlation (r^2^ = 0.68). The same analysis on only NAION TD pairings resulted in censoring thresholds of 19 dB for stimIII and 22 dB for stimV, and the resultant line of best fit had a slope of 0.67 with a similar correlation (r^2^ = 0.67).

## Discussion

We found that censoring and converting sensitivities to TDs enables direct comparison between stimIII and stimV. The average difference for censored and uncensored data is marginal, but the retest variability is markedly reduced using censoring. Determining an optimal censoring threshold for VFs for each stimulus improves the comparison between stimuli particularly in VFs with regions of major sensitivity loss.

We determined that the optimal censoring thresholds for stimIII and stimV are 21 dB and 24 dB for this data set of two different optic nerve disorders. These thresholds provided the lowest mean differences and low standard deviations, and are both within a reasonable range, allowing for better comparison between eyes with severely depressed VFs. These thresholds are similar to those calculated for each disease separately, and the lines of best fit for the combined and single disease computations have similar slopes and r² values. Therefore, our calculated overall thresholds of 21 dB and 24 dB for their respective stimuli may be applicable across a broader range of optic nerve diseases

A paired t-test revealed a small, but significant, average difference between TD pairings. However, this number is very close to zero, and considering same-day testing variability, this number has minimal clinical relevance. Prior studies have shown that the inter-test variability for specific points range from 1.3 dB - 3.0 dB for stimIII and 1.2 dB - 2.0 dB for stimV.^7^ Thus, the two stimuli should be largely interchangeable in clinical practice if censoring and age corrected TDs are used. This allows clinicians to switch from a test using one stimulus size to another without clinically meaningful discrepancies affecting clinical decisions. For NAION and glaucoma, the average TD difference and standard deviation were very similar (as well as between censored and uncensored pairs), suggesting that our methodology may broadly apply for a variety of optic nerve disorders.

The line of best fit for TD pairings closely followed the line of unity. Coupled with its high r^2^ value, suggests there is strong agreement between stimIII and stimV across different visual field severities. The variability in our uncensored data scatterplot highlights the necessity of censoring to maintain reliability and consistency.

This study has several clinical implications. Clinicians can choose between stimIII and stimV based on the degree of visual field damage without compromising diagnostic accuracy. We also demonstrated that patients undergoing regular VF testing with a mix of either stimuli will have reliable tracking of disease progression. Future research can leverage both stimIII and stimV within the same dataset, thereby eliminating the need to segregate data by Goldmann stimulus size. Our optimal censoring thresholds also improve the comparability of VFs not only within the same patient but across different patients as well in the setting of severe vision loss, aiding in better disease management.

Our study had limitations typical of retrospective analyses. First, widespread population data sets for VFs performed using stimulus V are lacking and are not included in the current Humphrey perimeters. Differences in testing protocols or equipment settings between the two datasets may have introduced variability that would affect the comparability of stimIII and stimV results, particularly due to the presence of multiple testing sites in both groups. Of course, having NAION VFs performed at 80 sites supports the potential for real world applicability of our method. We also only tested VFs of participants with NAION and glaucoma with substantial damage so we had relatively fewer data points near a TD of 0 and above.

Our study shows that usage of normative databases for stimIII and stimV, in conjunction with censoring, allows for the direct comparison between VFs using either of the two stimuli. Future directions may include validating these findings across diverse patient populations, extending the comparison to include other stimuli types, or comparing visual field patterns between the stimuli with methods such as archetypal analysis.

### Characteristics of Study Participants

**Table 1.** Demographic information for participants with glaucoma and non-arteritic anterior ischemic optic neuropathy.

|  | # VFs | Age (years) | % Male | % Right eye |
| --- | --- | --- | --- | --- |
| **Glaucoma**<br>**(n = 120)** | **1053** | **67.8 ± 9.3** | **39%** | **53%** |
| **NAION**<br>**(n = 586)** | **939** | **61.3 ± 7.7** | **69.3%** | **51%** |

### Mean TD Differences Between StimIII and StimV Pairs Across Censoring Thresholds

**Figure 1.**
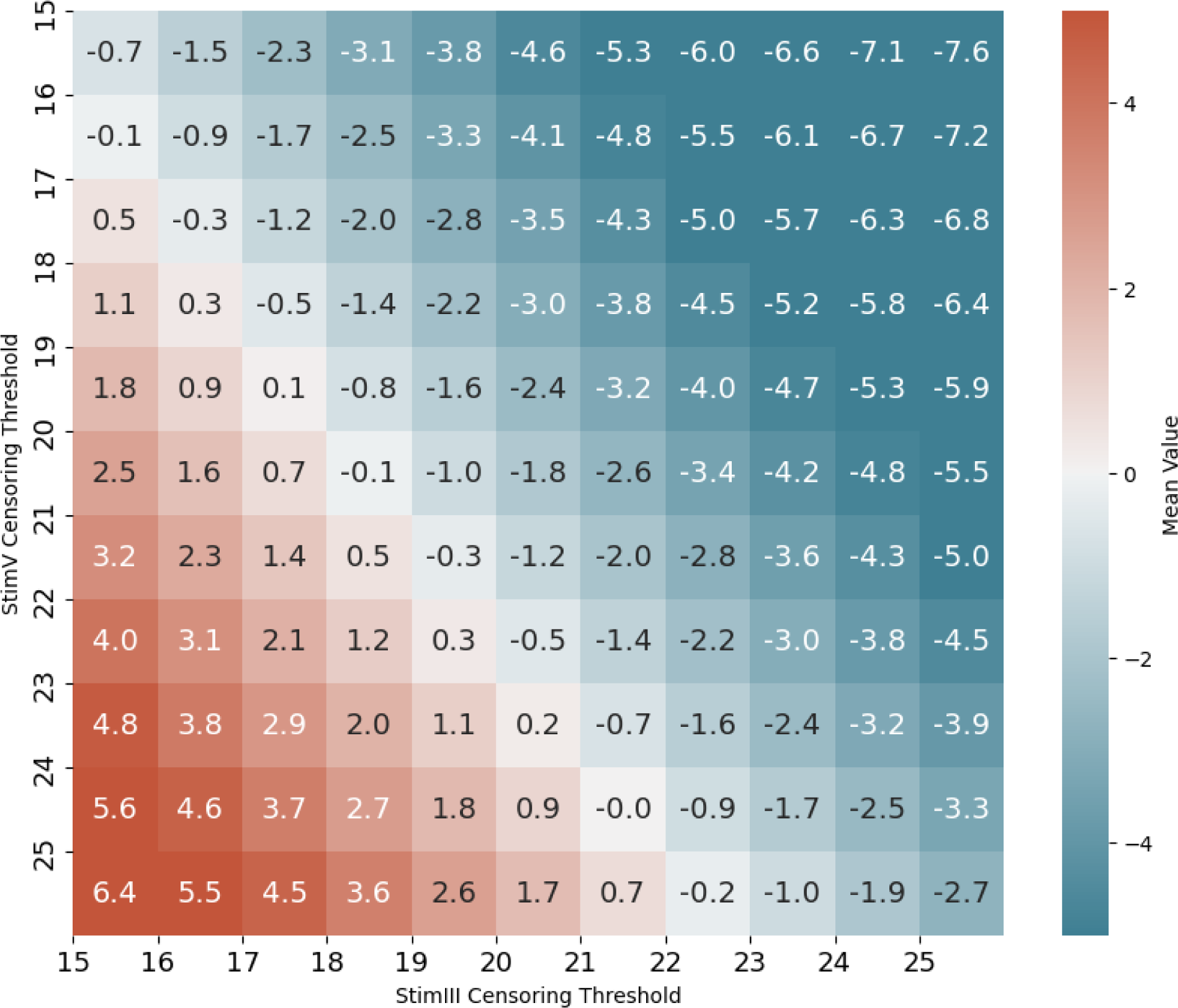
Heatmap depicting the average total deviation difference between pairs of stimulus size III and stimulus size V points across varying censoring thresholds where at least one stimulus was censored. The optimal censoring thresholds were identified at 21 dB for stimIII and 24 dB for stimV.

### Pointwise TD Differences Between StimV and StimIII

**Figure 2.**
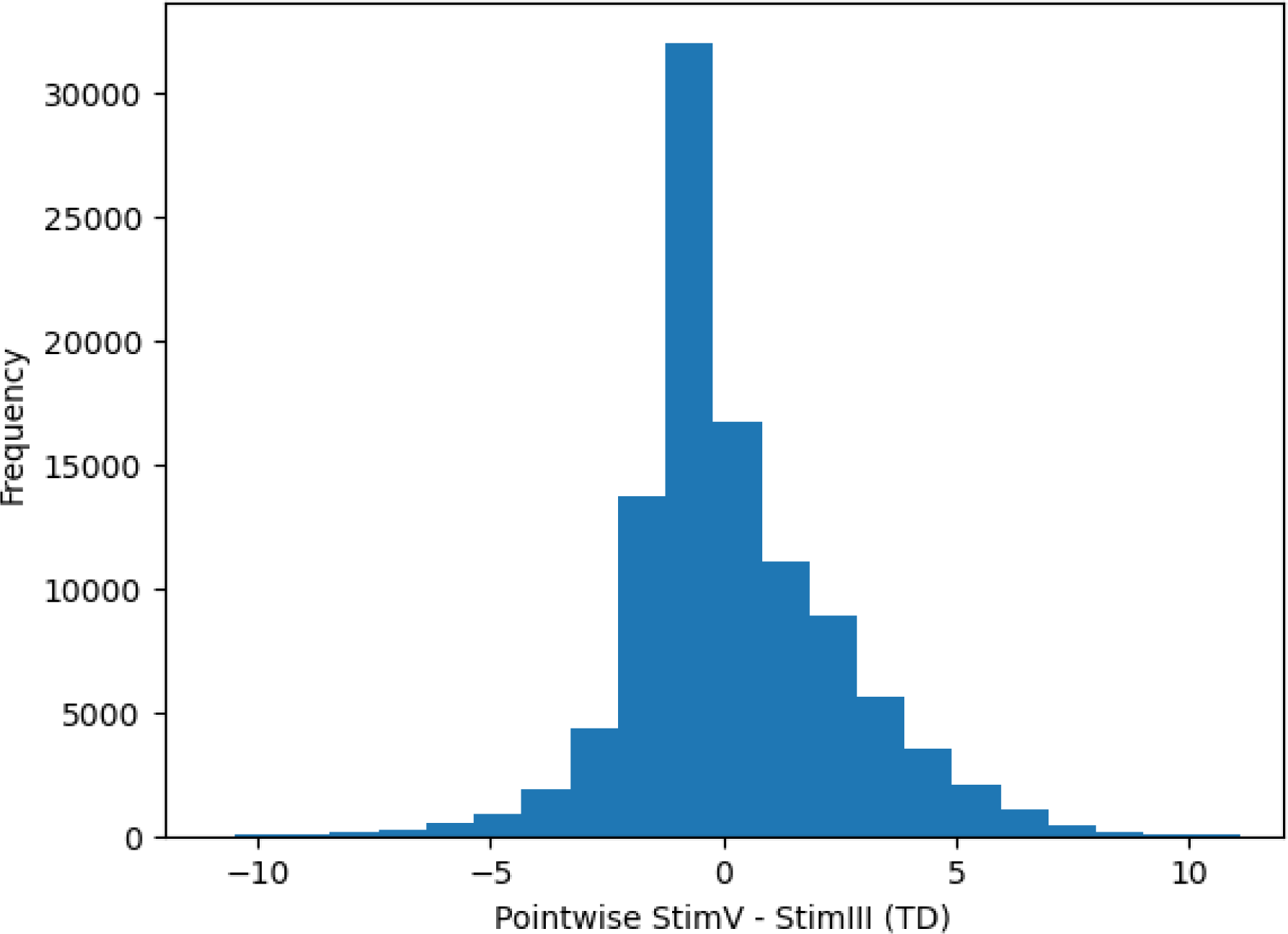
Histogram illustrating the distribution of pointwise total deviation differences between stimulus size V and stimulus size III visual fields. The histogram is centered around 0, indicating comparable differences between the two stimuli across the tested points.

### Scatterplot of TD Pairs Between StimIII and StimV

**Figure 3.**
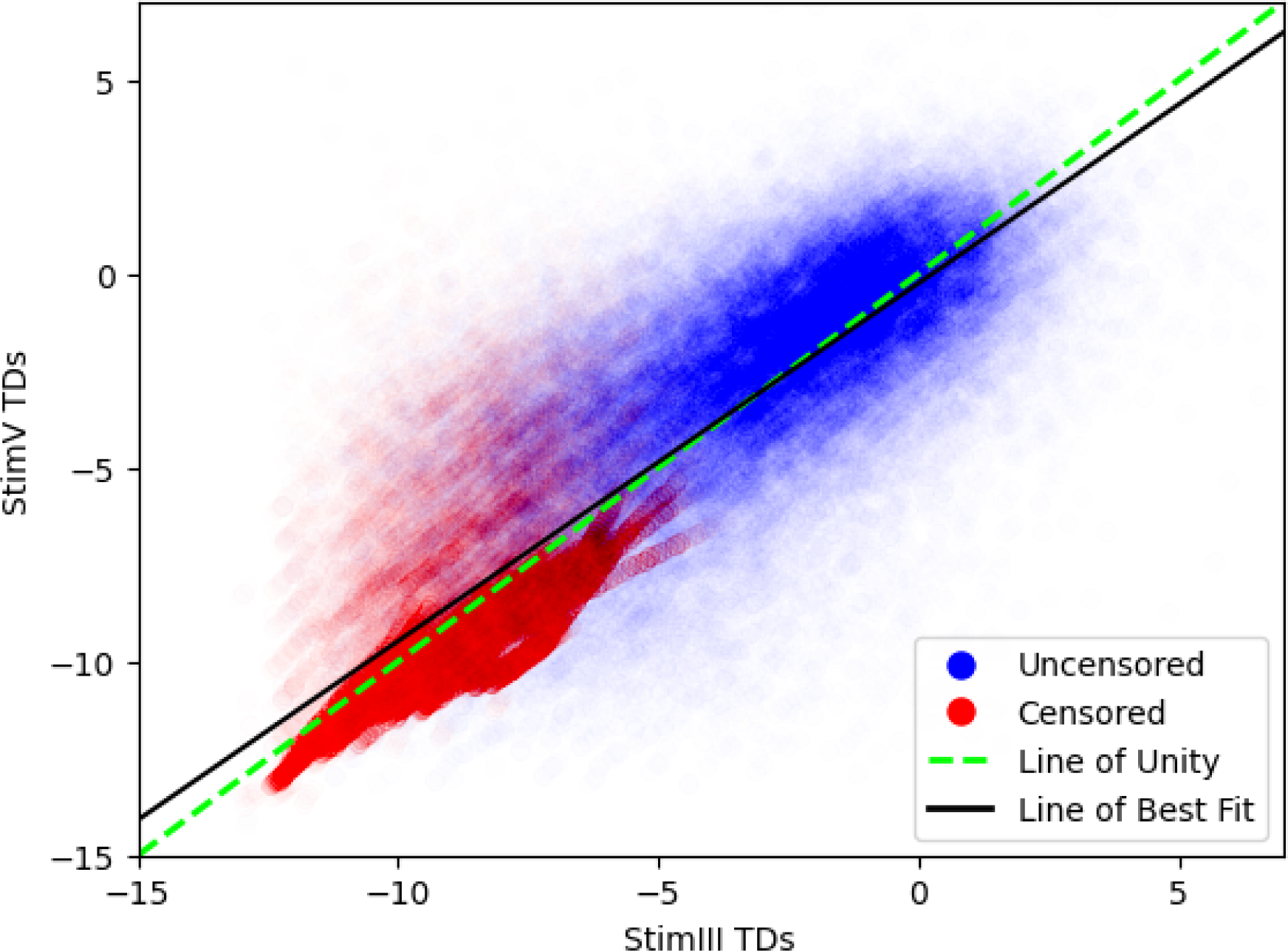
Pairs of total deviation values from stimulus size III and stimulus size V visual fields. Data points are colored based on whether at least one of the pair is censored. The dotted green line represents the line of unity (y = x). The black line of best fit (y = −0.24 + 0.92x) demonstrates a strong linear relationship between the two stimuli (r^2^ = 0.70). Data points are made transparent for improved visibility and to illustrate relative density.

### Scatterplot of Uncensored TD Pairs Between StimIII and StimV

**Figure 4.**
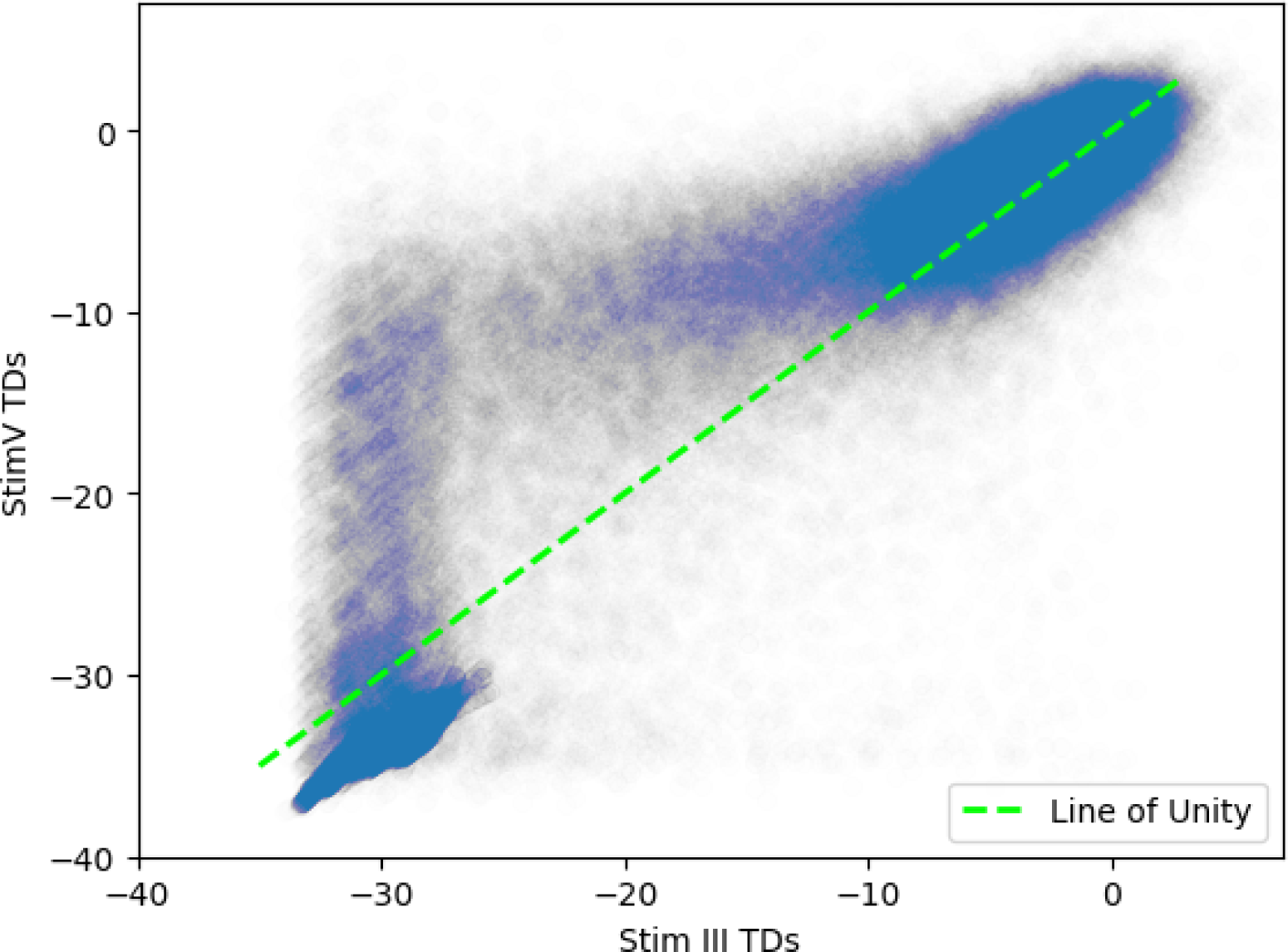
Pairs of uncensored total deviation values from stimulus size III and stimulus size V visual fields. The dotted green line represents the line of unity (y = x). Beyond a certain threshold, points increasingly deviate from the line of unity, highlighting point variability in uncensored measurements. Data points are made transparent for improved visibility and to illustrate their relative density.

## Data Availability

The data that support the findings of this study are not publicly available due to privacy and ethical restrictions. Patient information and medical records contain sensitive personal health information that cannot be shared publicly to protect patient confidentiality and comply with ethical guidelines.

---

**Supplemental Figure 1.**
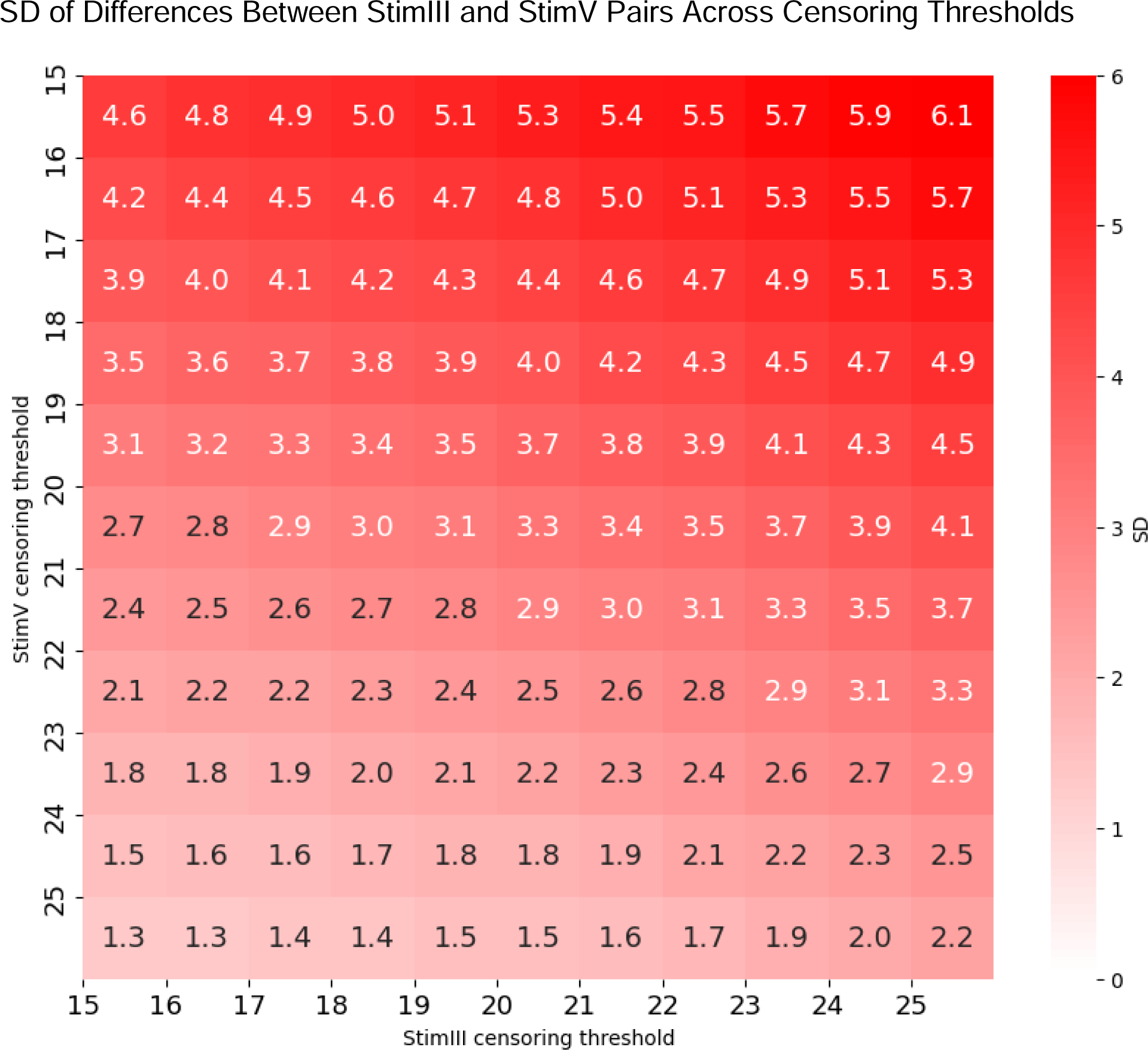
Heatmap depicting the standard deviation of total deviation differences between pairs of stimulus size III and stimulus size V points across varying censoring thresholds where at least one stimulus was censored. For the optimal censoring threshold of 21 dB for stimulus size III and 24 for stimulus size V, the standard deviation is 1.9 dB.

